# Preliminary diagnostic performance of the VIDAS® TB-IGRA for the detection of *Mycobacterium tuberculosis* infection and disease

**DOI:** 10.1101/2022.04.01.22271763

**Authors:** Serge Diagbouga, Arthur D. Djibougou, Camille Pease, Ariana Alcaide, Audrey Berthoux, Natalie Bruiners, Daniela Maria Cirillo, Ardjouma Combary, Nadine Falchero, Deborah Handler, Antoinette Kabore, Alfred Lardizabal, Amanda Lopes, Marissa Loubet, Philippe Manivet, Clemence Margain, Valerie Meunier, Faiza Mougari, Alberta Onyuka, Sophie Rivoiron, Tani Sagna, Mathilde Sanvert, Leon Sawadogo, Jacques Simpore, Emmanuelle Cambau, Maria Laura Gennaro

## Abstract

**Importance:** Accurate diagnosis of tuberculosis (TB) infection can be achieved with interferon-γ release assays. Their performance can be improved by utilizing fully automated, single-patient formats.

**Objective:** Establish clinical thresholds for a new interferon-γ release assay, the VIDAS® TB-IGRA, and compare diagnostic performance in detecting tuberculosis infection and disease with the established QuantiFERON-TB Gold Plus (QFT-Plus).

**Design:** Preliminary diagnostic performance study (October 2^nd^, 2019–February 20^th^, 2020). Setting: Multicenter.

**Participants:** Participants were divided into TB disease, high-risk, and low-risk populations. The confirmed TB disease population included 107 patients. The high-risk population included 162 individuals with flagged risk factors on a questionnaire but without objective clinical confirmation of TB. The Low-risk population included 117 healthy blood donors from the French National Blood Bank.

**Exposures:** Tuberculosis.

**Main Outcomes and Measures:** Positive and negative percent agreement (PPA, NPA) were determined between the VIDAS® TB-IGRA and QFT-Plus. In the TB disease population, sensitivity was also measured against bacterial culture and PCR.

**Results:** The VIDAS^®^ TB-IGRA produced fewer indeterminate results than the QFT-Plus (1/107 vs. 23/107) in the TB disease population. One analysis included indeterminate results as false negatives (94 positives and 10 false negatives vs. 56 positives and 48 false negatives), and the VIDAS^®^ TB-IGRA exhibited higher sensitivity than the QFT-Plus (90.4% vs. 53.8%) (p<0.0001). Another analysis excluded indeterminate results (76 positives and 4 false negatives vs. 55 positives and 25 false negatives), and the VIDAS® TB-IGRA again exhibited higher sensitivity than the QFT-Plus (95.0% vs. 68.8%) (p<0.0001). A 98.2% PPA was calculated between the two tests with this dataset.

In the high-risk population, the VIDAS® TB-IGRA exhibited a strong PPA (94.4%) with the QFT-Plus. However, a lower than expected NPA was observed (85.2%). In the low-risk population, the VIDAS® TB-IGRA demonstrated high specificity (94.9%) and a strong NPA (98.2%) with the QFT-Plus.

**Conclusions and Relevance:** The fully automated VIDAS^®^ TB-IGRA is a promising diagnostic test for both TB infection and disease. It exhibits higher sensitivity while maintaining specificity and produces fewer indeterminate interpretations. Its easy-to-use, single-patient format may lead to increased TB testing to help with the worldwide eradication of the disease.

**KEY POINTS:** *Question:* What is the diagnostic performance of the VIDAS® TB-IGRA in detecting tuberculosis infection and disease?

*Findings:* The VIDAS^®^ TB-IGRA exhibited high sensitivity in individuals with tuberculosis disease (90.4– 95.0%), high specificity in healthy blood donors (94.9%), a high positive percent agreement (PPA) in individuals with a high risk of tuberculosis infection (94.4%), and it produced a low number of indeterminate results (1/386).

*Meaning:* The VIDAS^®^ TB-IGRA is a promising diagnostic test for both tuberculosis infection and disease.

## INTRODUCTION

Tuberculosis (TB) is an infection caused by a bacterial pathogen (*Mycobacterium tuberculosis*) that most often affects the lungs. Globally, it is one of the leading causes of death from a single infectious agent^1^. According to the World Health Organization (WHO), 1.4 million individuals died from TB and approximately 10 million became ill with the disease in 2019. Furthermore, approximately one-fourth of the global population is estimated to have TB infection. Individuals with TB infection do not exhibit symptoms and cannot transmit the disease; however, they have a 5–15% risk of becoming ill and potentially spreading the disease within their lifetime^2^. TB is preventable and treatable; however, the number of cases being properly diagnosed and treated remains low^1^. Therefore, the quick and accurate detection of both *M. tuberculosis* infection and disease is essential to adequately control this pandemic.

Currently used diagnostic methods for TB infection and disease include bacterial culture, PCR, chest radiography, the tuberculin skin test (TST), and interferon-γ release assays (IGRAs). Bacterial culture is the current gold standard for the diagnosis of TB disease, followed by PCR, whereas the TST and IGRAs are the most commonly used methods to diagnose TB infection. The TST has been available and widely used since the 1900s. It uses a heat-killed purified protein derivative from *M. tuberculosis* cultures, which shares components with other mycobacteria. Therefore, the TST cannot distinguish between *M. tuberculosis* infection and reactivity due to previous vaccination with the Bacillus Calmette-Guerin (BCG) vaccine or infections caused by non-tuberculous mycobacteria (NTM)^1,3-5^.

Since 2001, the development of IGRAs has provided many benefits over the TST for the detection of TB infection. The QuantiFERON-TB Gold Plus (QFT-Plus) and T-SPOT-TB have been the most widely used, FDA-approved IGRAs that use synthetic peptides derived from M. tuberculosis-specific early secreted antigenic target 6 (ESAT-6) and culture filtrate protein 10 (CFP-10). These tests are used to detect *ex vivo* reactivity specifically associated with *M. tuberculosis* infection. False-positive results due to BCG cross-reaction are very low with these IGRAs because the synthetic peptides used are absent in BCG vaccine strains and most pathogenic NTM strains. Additionally, these IGRAs only require a single patient visit and results are available within 24 hours of testing, unlike the TST that requires a second visit within 48–72 hours^4-6^.

Although the development of IGRAs represents a huge breakthrough in diagnosing TB infection, many drawbacks remain. Currently available IGRAs involve multiple cumbersome manual steps, which can increase the likelihood of human error and introduce variability in the execution of the preanalytical steps, potentially leading to erroneous results and interpretations^6-11^. Additionally, these IGRAs are typically performed as batch tests that require several patient samples per microplate, which may delay results by several days^9,10^.

The newly developed, fully automated VIDAS® TB-IGRA offers an improved method for the detection of both TB infection and disease. The fully automated system uses the commercially available VIDAS3 platform to quickly deliver results in one easy step, thus eliminating the multiple steps required by other IGRAs, decreasing the occurrence of human (or operator) error, and enhancing robustness with lower variability in results. Furthermore, the process is performed in a single-patient format, which allows for a quick (within 17 hours) turn-around time.

In the present study, we evaluated and compared the performance of the newly designed VIDAS® TB-IGRA with that of the QFT-Plus to detect both TB infection and disease in three patient populations, including individuals with confirmed TB disease as well as populations with a high or low risk of TB infection. Our data demonstrate an improved detection capacity with maintained specificity for the optimized VIDAS® TB-IGRA, thus suggesting its value in diagnosing both TB infection and disease.

## METHODS

### Participants

Participants were divided into TB disease, high-risk, and low-risk populations according to the eligibility criteria outlined in Supplemental Figure 1. TB disease was determined by bacterial culture or PCR. All study participants in the TB disease group were ≥2 years of age and were HIV negative.

The high-risk population consisted of patients who had an increased risk of *M. tuberculosis* exposure or infection based on a patient questionnaire. Individuals were considered to be at a high risk of TB infection if they lived with someone with TB disease, lived in or emigrated from a country with a high prevalence of TB disease, spent more than 1 month in an area with a high prevalence of TB infection, belonged to a group with high TB transmission rates (e.g., homeless, incarcerated, or previously incarcerated individuals, intravenous drug users), abused drugs and/or alcohol, worked or resided at facilities with high-risk individuals (e.g., hospitals, homeless shelters, correctional facilities, nursing homes, HIV patient residences), were children or adolescents exposed to high-risk adults, or were chronically immunocompromised or on immunosuppressive treatments. All study participants of the high-risk population were ≥2 years of age.

The low-risk population consisted of individuals who were previously questioned concerning their history of TB and accepted by the French National Blood Bank as healthy blood donors. All study participants of the low-risk population were ≥18 years of age.

Any individuals who had received >15 days treatment for ongoing TB disease, received anti-TNF-α treatment within the past 3 months, who had a positive TST within the past 12 weeks, who had been diagnosed with an NTM infection, or who had been diagnosed with HIV at the time of inclusion or 1 month prior were excluded from the study. Pregnant women were also excluded from the study. Additional patient information was collected for data monitoring and follow-up (Supplemental Table 1). Written informed consent was obtained from all study participants.

### Sample collection and diagnostic testing

Two whole-blood samples were collected from each participant into 4-mL lithium heparin blood collection tubes. One sample was used for the VIDAS^®^ TB-IGRA test, and the other was used for the QFT-Plus test.

Samples were obtained and tested from October 2^nd^, 2019 to February 20^th^, 2020 at the four following sites: Institut de Recherche en Sciences de la Santé (IRSS), Ouagadougou, Burkina Faso; Public Health Research Institute, Rutgers New Jersey Medical School, Newark, NJ, USA; Hôpital Lariboisière AP-HP/Assistance Publique-Hopitaux de Paris, France; and samples obtained from the French National Blood Bank (EFS) were tested at bioMerieux Marcy l’etoile, Rhone-Alpes, France. Samples were routinely stored either at 2–8°C for ≤32 hours or at 18–25°C for ≤6 hours prior to testing.

The VIDAS^®^ TB-IGRA and QFT-Plus diagnostic tests were simultaneously performed for the samples, according to the approved VIDAS^®^ TB-IGRA protocol (see below) and the QFT-Plus manufacturer’s instructions. For the VIDAS® TB-IGRA, each sample was automatically subset into a negative control (NIL) sample, an antigen response sample (AG-NIL), and a mitogenic response positive control (MIT-NIL) sample. For the QFT-Plus, each sample was subset into a negative control (Nil) sample, two antigen response samples (TB1 and TB2), and a mitogenic response positive control (Mitogen) sample. Sample interferon-γ (IFN-γ) values were used to obtain a positive, negative, or indeterminate interpretation based on the established thresholds for each method.

Ethics committee approval was obtained for all study protocols (IRSS: 25 A012-2019/CEIRES, Rutgers University: IRB00000607, Hôpital Lariboisière AP-HP/Assistance Publique-Hopitaux de Paris: BB-0033-00064, EFS: AC-2017-2958).

### VIDAS^®^ TB-IGRA

The VIDAS^®^ TB-IGRA test was performed using the supplied reagents and the VIDAS3 system. Each whole-blood sample was tested using three VIDAS^®^ reagent strips and three solid phase receptacles (SPRs). The samples were loaded into the instrument with three stimulation reagents (AG, antigen; MIT, positive control; NIL, negative control). Reagent transfers were performed by the VIDAS3 automatic pipetting unit (APU). The samples were homogenized and incubated with the individual stimulation reagents for 16 hours at 37°C in the strips. Next, the automated system performed an enzyme-linked immunofluorescent assay (ELFA) to quantify the sample IFN-γ concentrations. The final results were interpreted using internal calculation tools and previously established thresholds (Figure 1 and Table 1).

**Figure 1.**
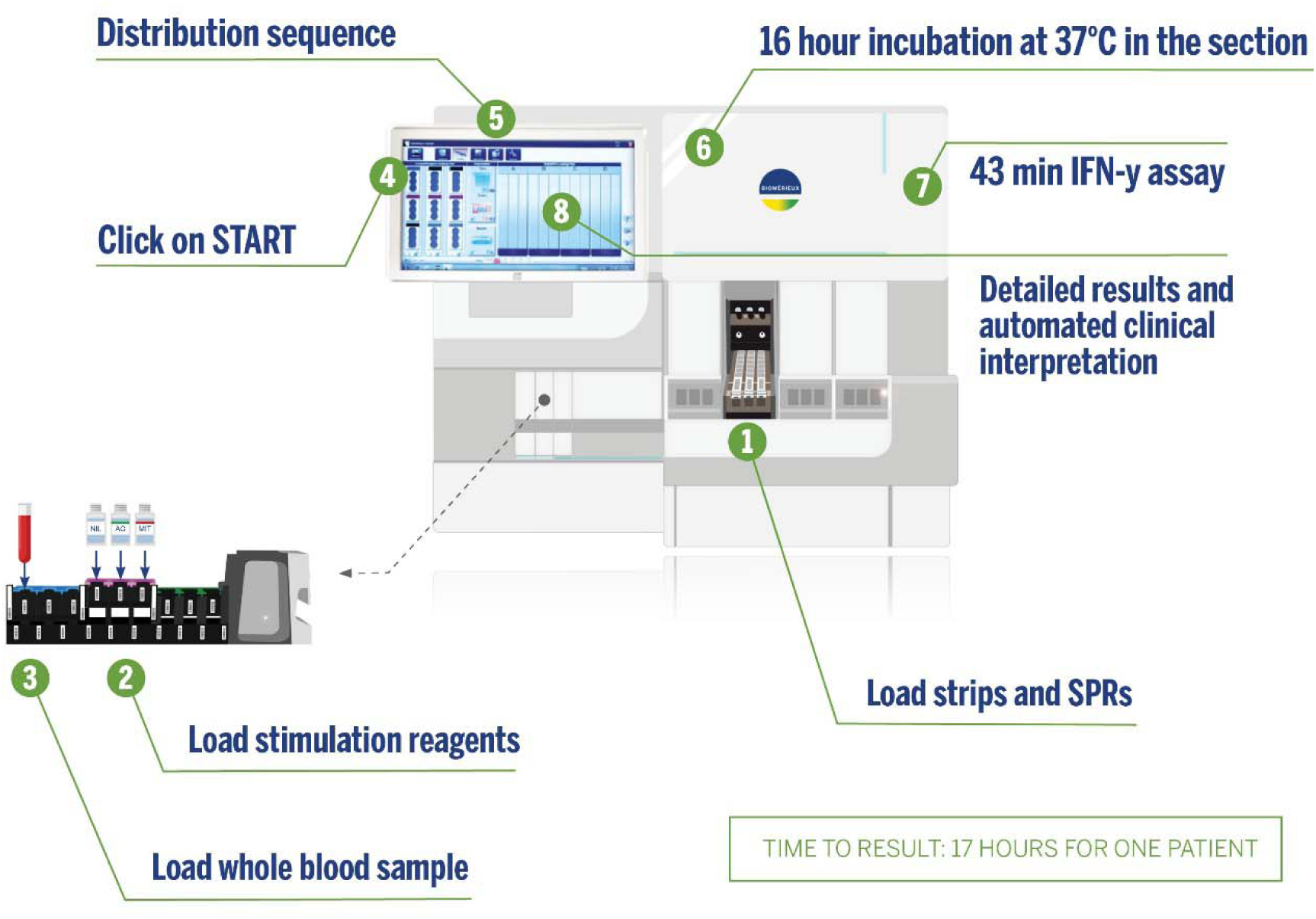
VIDAS® TB-IGRA workflow.

**Table 1.** VIDAS® TB-IGRA threshold values.

| NIL (IU/mL) | AG-NIL (IU/mL) | MIT-NIL (IU/mL) | Interpretation | Clinical significance |
| --- | --- | --- | --- | --- |
| <6.4 | $\geq 0.35$ and $\geq 25\%$ NIL | Any | Positive | *M. tuberculosis* infection likely |
| | <0.35<br>OR<br>$\geq 0.35$ and <25% NIL | $\geq 1.1$ | Negative | *M. tuberculosis* infection NOT likely |
| | <0.35<br>OR<br>$\geq 0.35$ and <25% NIL | <1.1 | Indeterminate<br>MIT Low | Likelihood of *M. tuberculosis* infection cannot be determined |
| $\geq 6.4$ | Any | | Indeterminate<br>NIL High | |

Briefly, AG-NIL IFN-γ concentrations ≥0.35 IU/mL indicated a positive result, whereas concentrations ≤0.35 IU/mL indicated a negative result. Results were considered indeterminate if MIT-NIL IFN-γ concentrations were <1.1 IU/mL or if NIL IFN-γ concentrations were ≥6.4 IU/mL.

### Statistical analyses

To establish statistical significance between the results of the QFT-Plus and VIDAS® TB-IGRA tests in the TB disease population, data were arranged into a contingency table and McNemar’s test was applied. A p-value <0.05 was considered statistically significant. Negative percent agreement (NPA) and positive percent agreement (PPA) were calculated to evaluate the performance of the VIDAS® TB-IGRA compared with that of the QFT-Plus.

## RESULTS

### TB disease population

In this study, a total of 107 patient samples from three locations (Ouagadougou, Burkina Faso, 101; Newark, NJ, USA, 4; and Paris, France, 2) were included in the TB disease population. TB disease was confirmed by bacterial culture or PCR. These confirmed TB disease cases were used to compare the diagnostic performance of the VIDAS® TB-IGRA and QFT-Plus tests. Three samples were excluded when calculating the PPA between the two tests and the sensitivity of the QFT-Plus because no QFT-Plus results were available.

In this TB disease population, only one indeterminate result was obtained with the VIDAS® TB-IGRA, whereas 23 were obtained with the QFT-Plus. Two statistical analyses were performed to estimate sensitivity using equivalent datasets between the two tests (Figure 2A).

**Figure 2.**
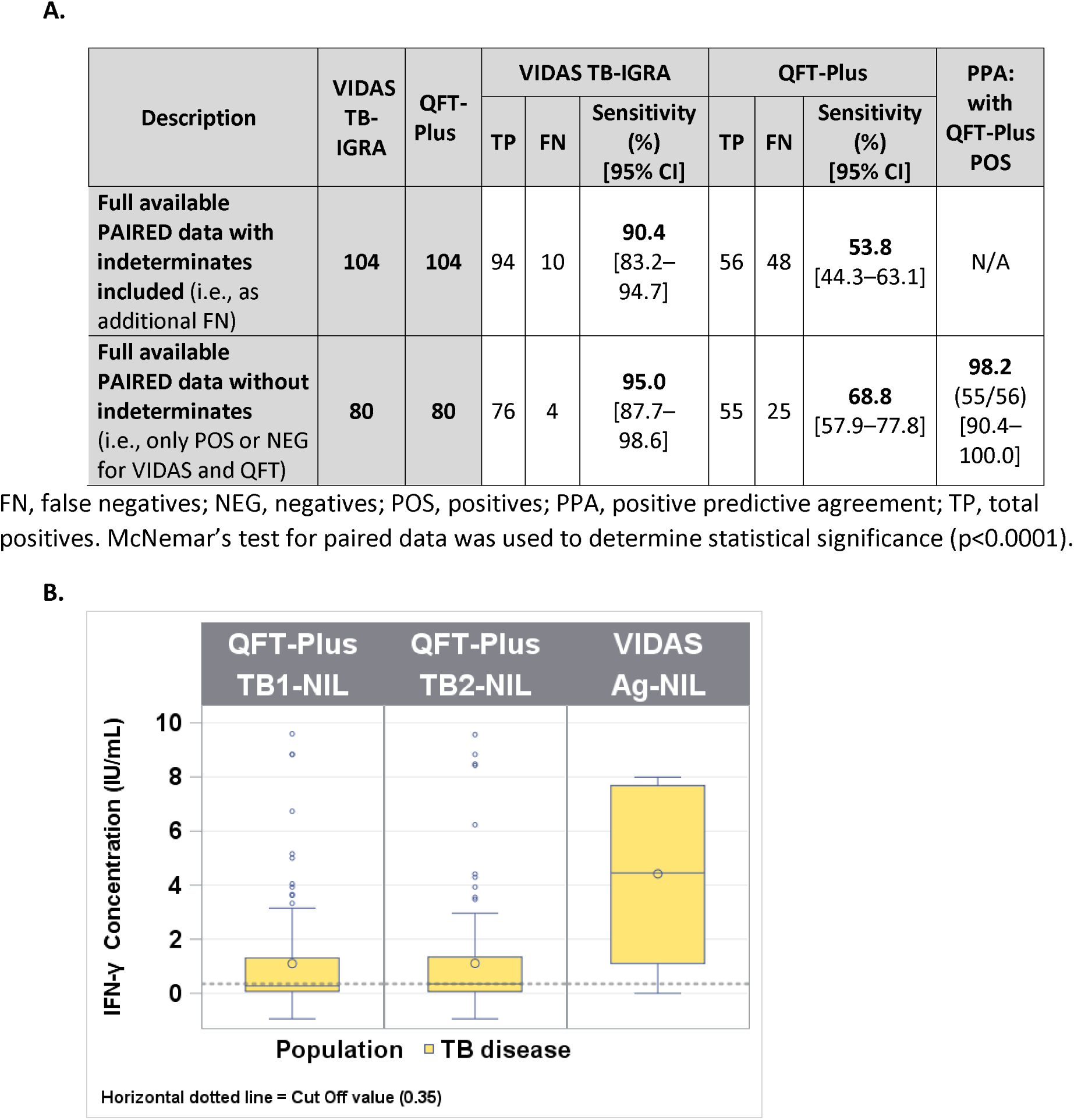
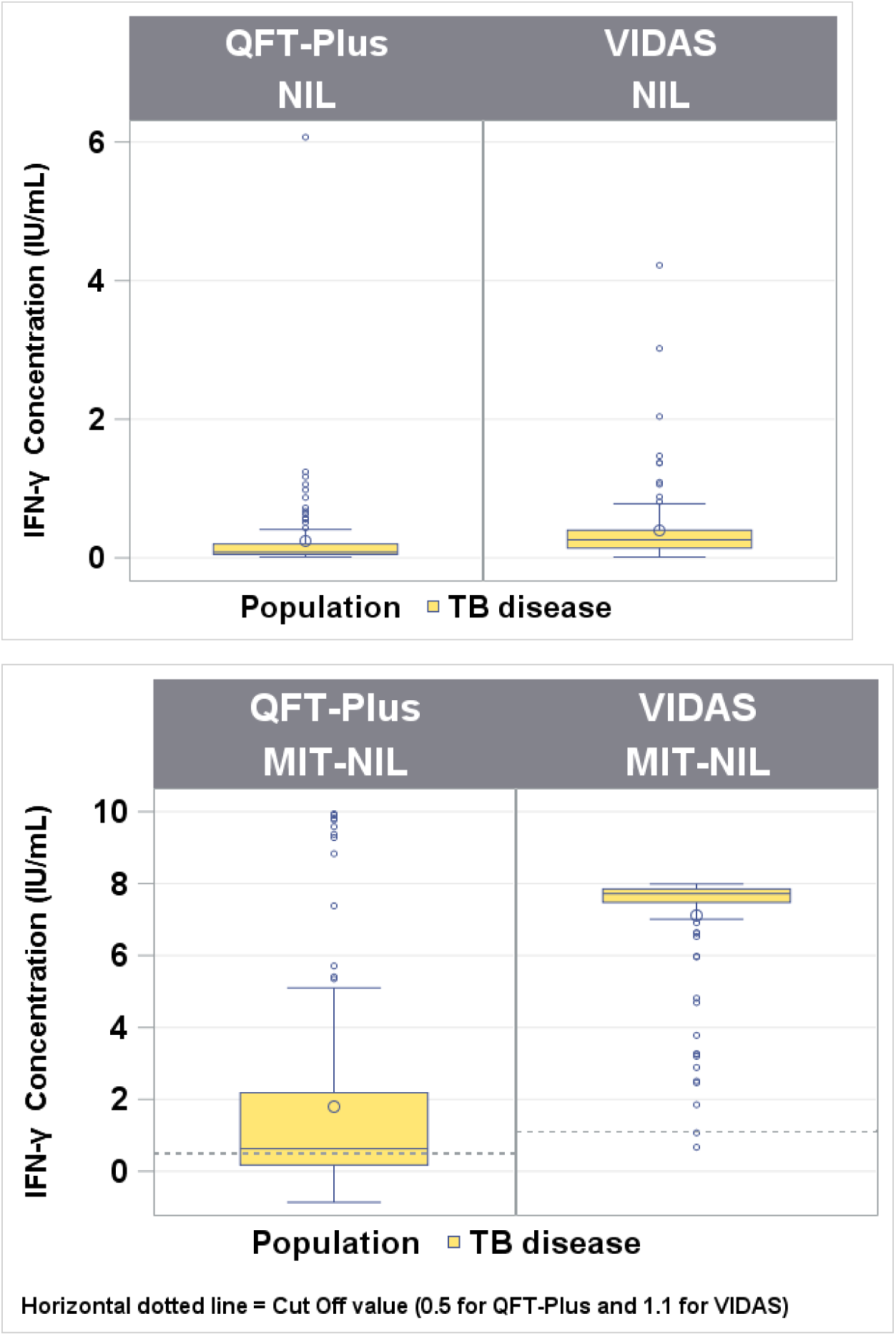
Comparison between the VIDAS® TB-IGRA and QFT-Plus for the tuberculosis disease population. **A.** Sample numbers, sensitivity, and positive predictive agreement (PPA). **B.** Patient sample interferon-γ (IFN-γ) concentrations for the VIDAS® TB-IGRA and QFT-Plus. Nil, QFT-Plus negative control; NIL, VIDAS® TB-IGRA negative control; Mitogen, QFT-Plus positive control; MIT-NIL, VIDAS® TB-IGRA positive control; TB1/TB2, QFT-Plus mitogenic response samples; AG-NIL, VIDAS® TB-IGRA mitogenic response sample. The dotted lines represent the sample threshold values for the respective assay. FN, false negatives; NEG, negatives; POS, positives; PPA, positive predictive agreement; TP, total positives. McNemar’s test for paired data was used to determine statistical significance (p<0.0001).

The first analysis used results from 104 patients and included the indeterminate results as false negatives (94 positives and 10 false negatives vs. 56 positives and 48 false negatives). In this analysis, the VIDAS® TB-IGRA exhibited a sensitivity of 90.4%, whereas the QFT-Plus exhibited a sensitivity of 53.8% (p<0.0001).

The second analysis excluded the indeterminate results (76 positives and 4 false negatives vs. 55 positives and 25 false negatives) and used results from 80 patients. In this case, the VIDAS® TB-IGRA exhibited a sensitivity of 95.0%, whereas the QFT-Plus exhibited a sensitivity of 68.8% (p<0.0001). Furthermore, a 98.2% PPA was calculated between the two tests with this dataset.

In the antigen response samples (AG-NIL vs. TB1/2-NIL) and positive control samples (MIT-NIL vs. Mitogen), the VIDAS® TB-IGRA detected higher IFN-γ values than the QFT-Plus for this population (Figure 2B). These higher values may have contributed to the improved detection capacity of the VIDAS® TB-IGRA.

Additional information was collected from Burkina Faso patients who presented indeterminate results with low IFN-γ values for the positive control samples. The number of CD4^+^ T cells was analyzed in these patient samples to determine whether these low responses were due to a low number of CD4^+^ T cells responding to the mitogenic stimulation. The VIDAS® TB-IGRA detected higher IFN-γ values in the AG-NIL and MIT-NIL samples compared with the QFT-Plus TB1/2 and Mitogen samples, regardless of the number of CD4^+^ T cells present (Supplemental Figure 2). Therefore, the differences observed between the two assays did not correlate with the number of CD4^+^ T cells. Many patients from the Burkina Faso location exhibited comorbidities and/or low body mass indexes, which may be an explanation for the low sensitivity of the QFT-Plus in these patients with TB disease (see Supplemental Results).

### High-risk population

A total of 162 patient samples from two locations (Ouagadougou, Burkina Faso, 90; Newark, NJ, USA, 72) were included in the high-risk population for TB infection. The Burkina Faso samples were all contact cases. In contrast, the Newark, NJ samples consisted of contact cases, immigrants from countries with a high prevalence of TB disease, and healthcare workers with an increased risk of exposure. Of these samples, 67 were determined as positive and 95 were determined as negative by the VIDAS® TB-IGRA, whereas 54 were determined as positive and 108 were determined as negative by the QFT-Plus. No indeterminate results were obtained with either diagnostic test.

We observed a strong PPA of 94.4% but a lower NPA of 85.2% (Figure 3A). Because the NPA was lower than expected, additional gradient analysis was conducted by introducing additional risk stratification in which individuals were categorized into subgroups according to their risk of *M. tuberculosis* exposure (Figure 3B). With this analysis, we observed a decrease in the NPA as the risk of *M. tuberculosis* exposure increased (Figure 3C and 3D).

**Figure 3.**
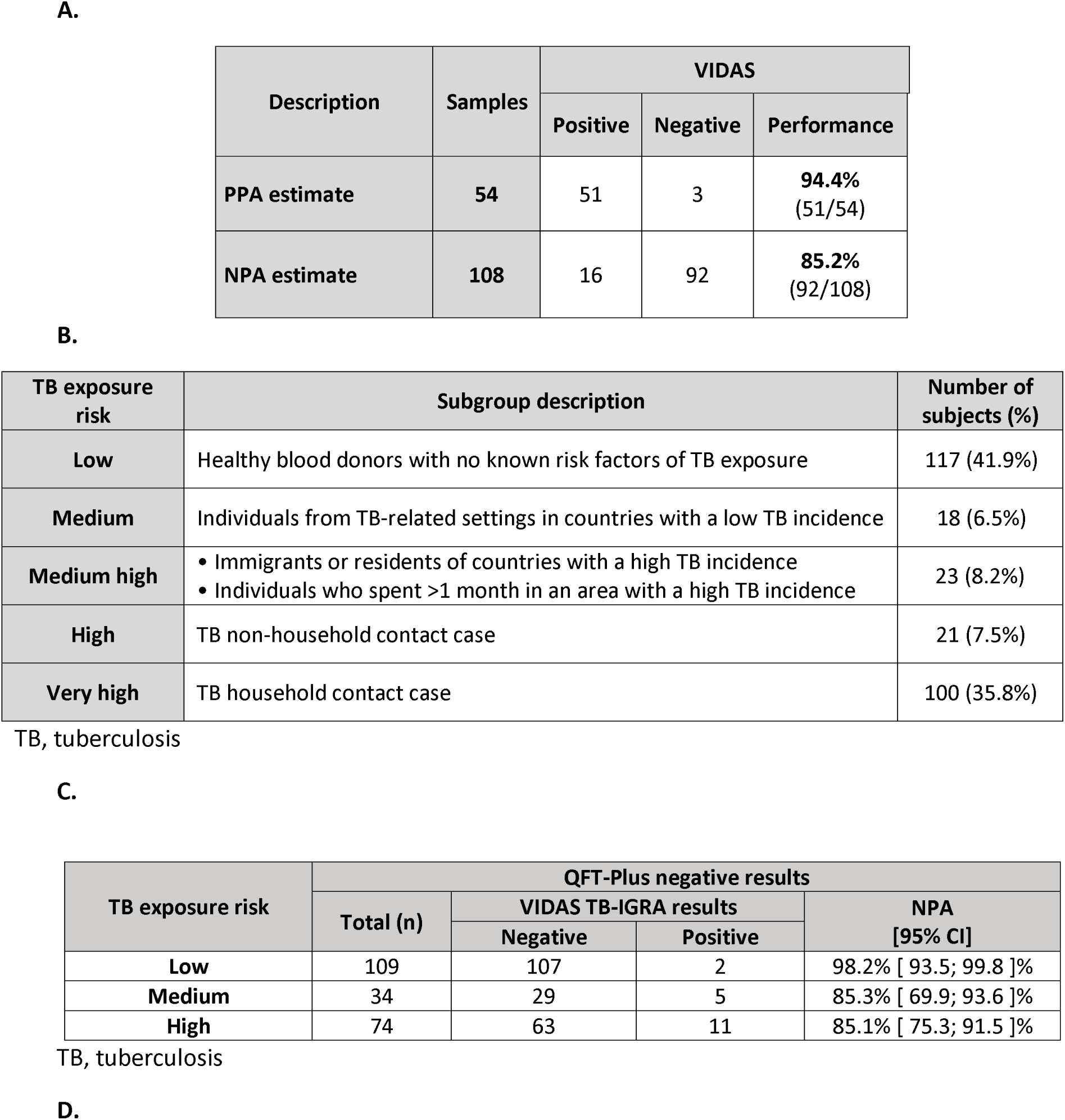
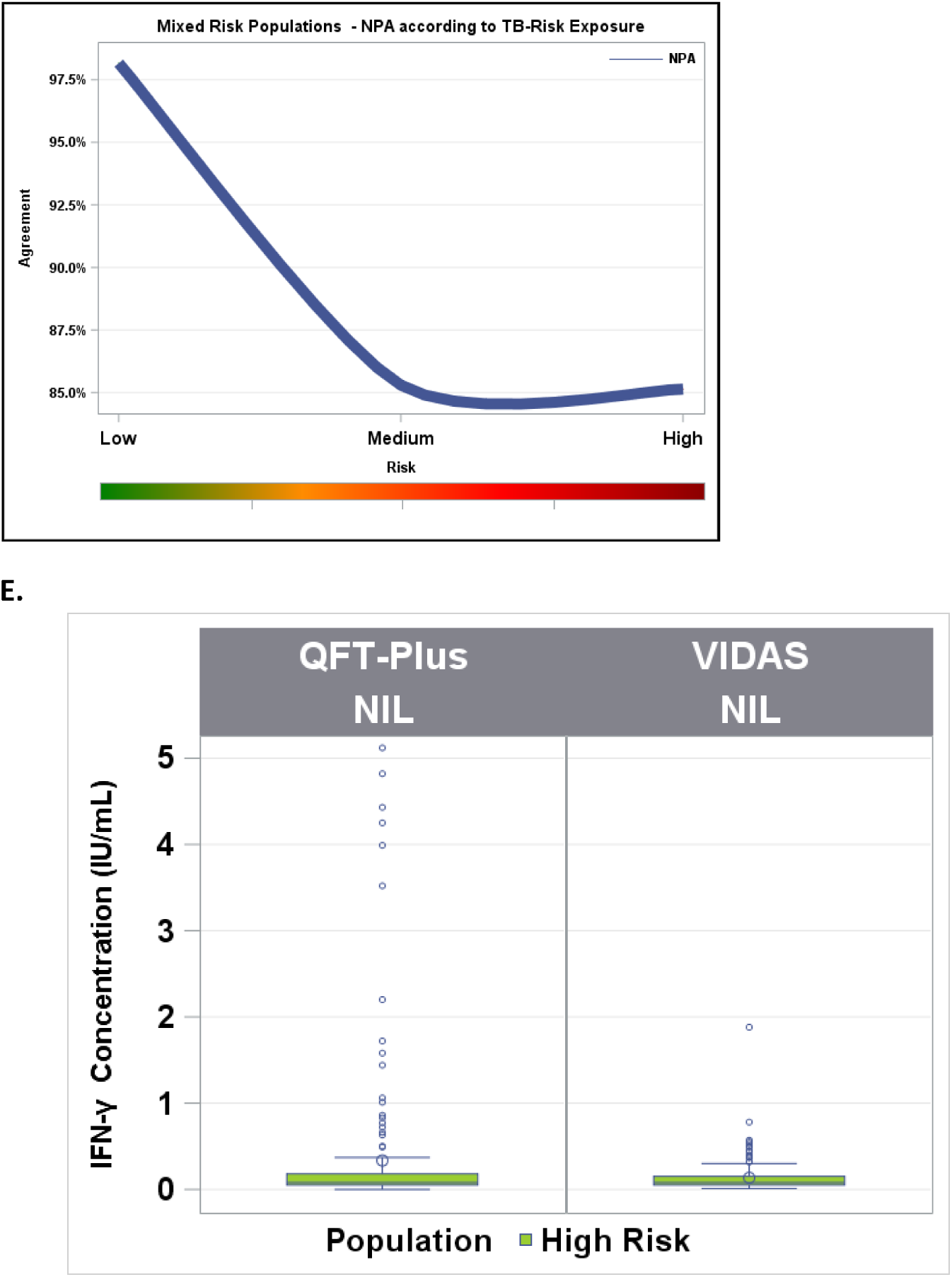
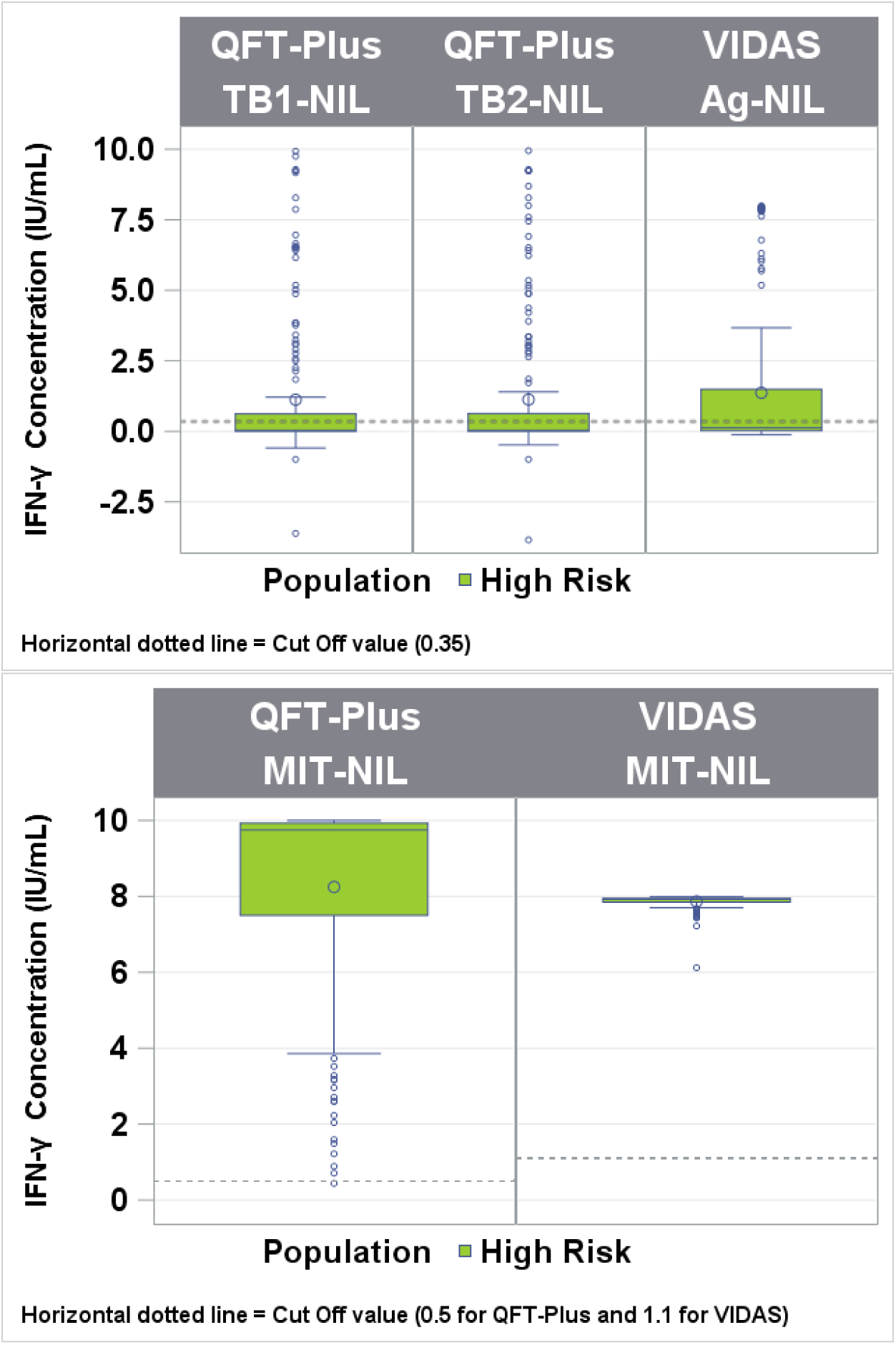
Comparison between the VIDAS^®^ TB-IGRA and QFT-Plus for the high-risk population. **A.** Sample numbers, positive predictive agreement (PPA), and negative predictive agreement (NPA). **B.** Subgroup stratification according to patient risk of tuberculosis exposure for gradient analysis. **C.** and **D.** NPA between the VIDAS^®^ TB-IGRA and QFT-Plus results according to patient risk of tuberculosis exposure. **E.** Patient sample interferon-γ (IFN-γ) concentrations for the VIDAS^®^ TB-IGRA and QFT-Plus. Nil, QFT-Plus negative control; NIL, VIDAS^®^ TB-IGRA negative control; Mitogen, QFT-Plus positive control; MIT-NIL, VIDAS® TB-IGRA positive control; TB1/TB2, QFT-Plus mitogenic response samples; AG-NIL, VIDAS^®^ TB-IGRA mitogenic response sample. The dotted lines represent the sample threshold values for the respective assay.

The distribution of the quantitative results showed that the VIDAS® TB-IGRA detected higher IFN-y values, resulting in the identification of more positive samples than the QFT-Plus (Figure 3E). This discrepancy was independently observed at the two locations that recruited the majority of the high-risk patients at a similar level (13.3% and 9.7% of the total results were discrepant in Burkina Faso and Newark, NJ, respectively). The discrepant results for this high-risk population are provided in Supplemental Table 2.

### Low-risk population

A total of 117 healthy blood donor samples from the EFS in the Rhone-Alpes Auvergne region of France were included in the low-risk population for TB infection. In this population, the VIDAS^®^ TB-IGRA detected six positive and 111 negative samples. Likewise, the QFT-Plus detected eight positive and 109 negative samples. No indeterminate results were obtained with either diagnostic test.

These healthy blood donors were from a country with very low prevalence of TB, and participants with any TB history were excluded from sample recruitment. Therefore, this population presented an extremely low risk of infection and was used to evaluate the specificity of the two tests. The VIDAS® TB-IGRA exhibited a specificity of 94.9%, and the QFT-Plus exhibited a similar specificity of 93.2% (Figure 4A).

**Figure 4.**
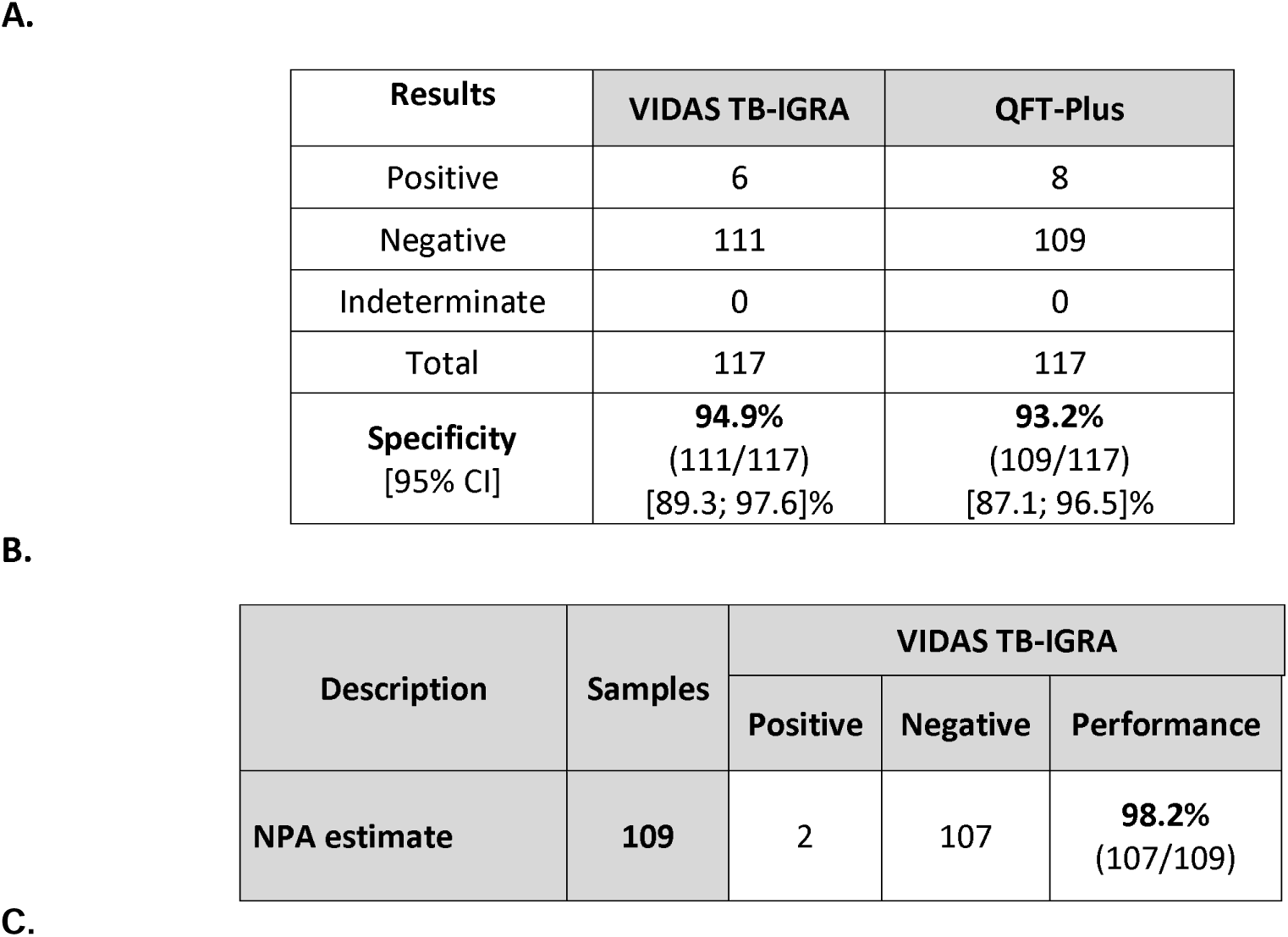
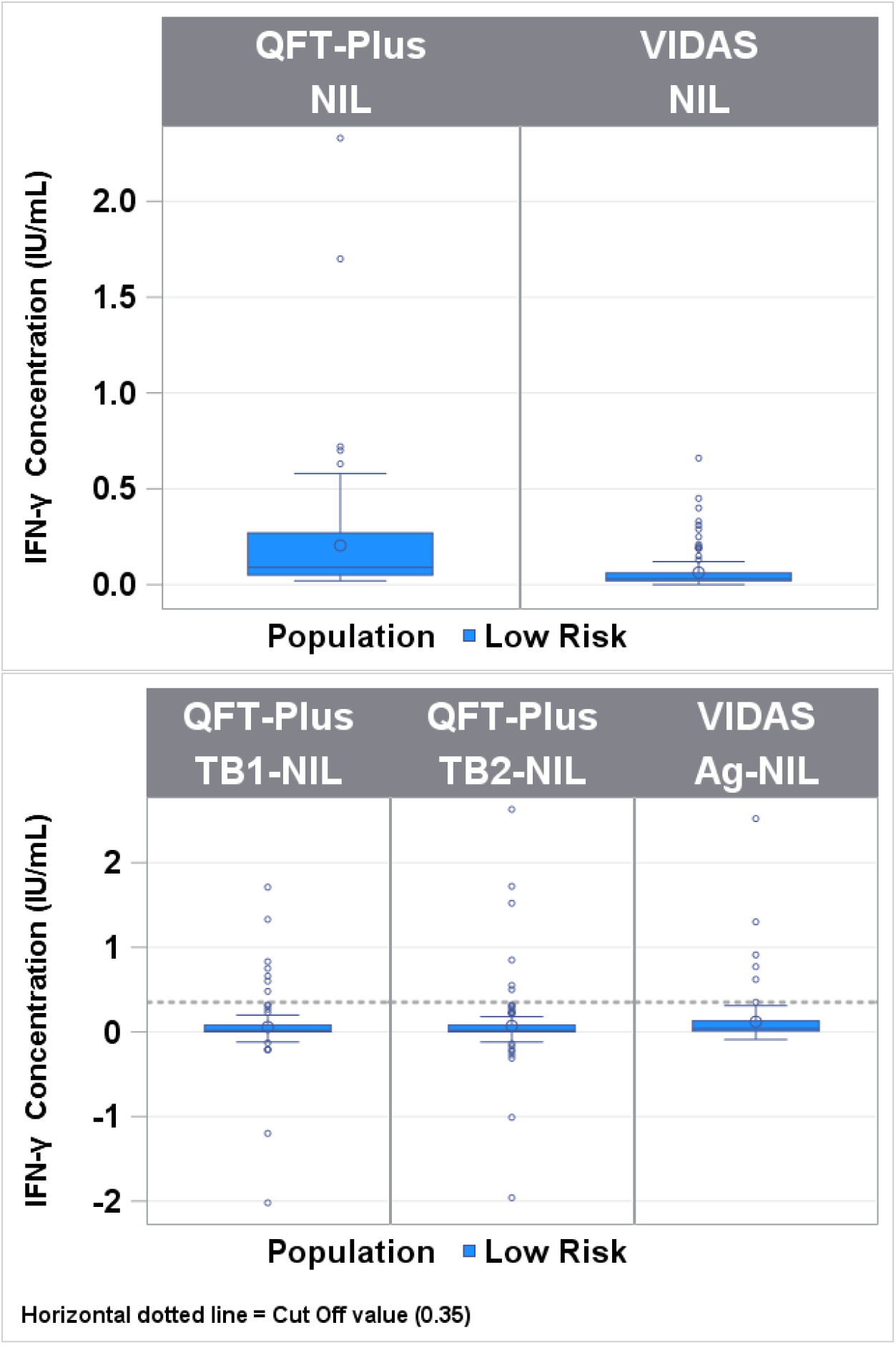
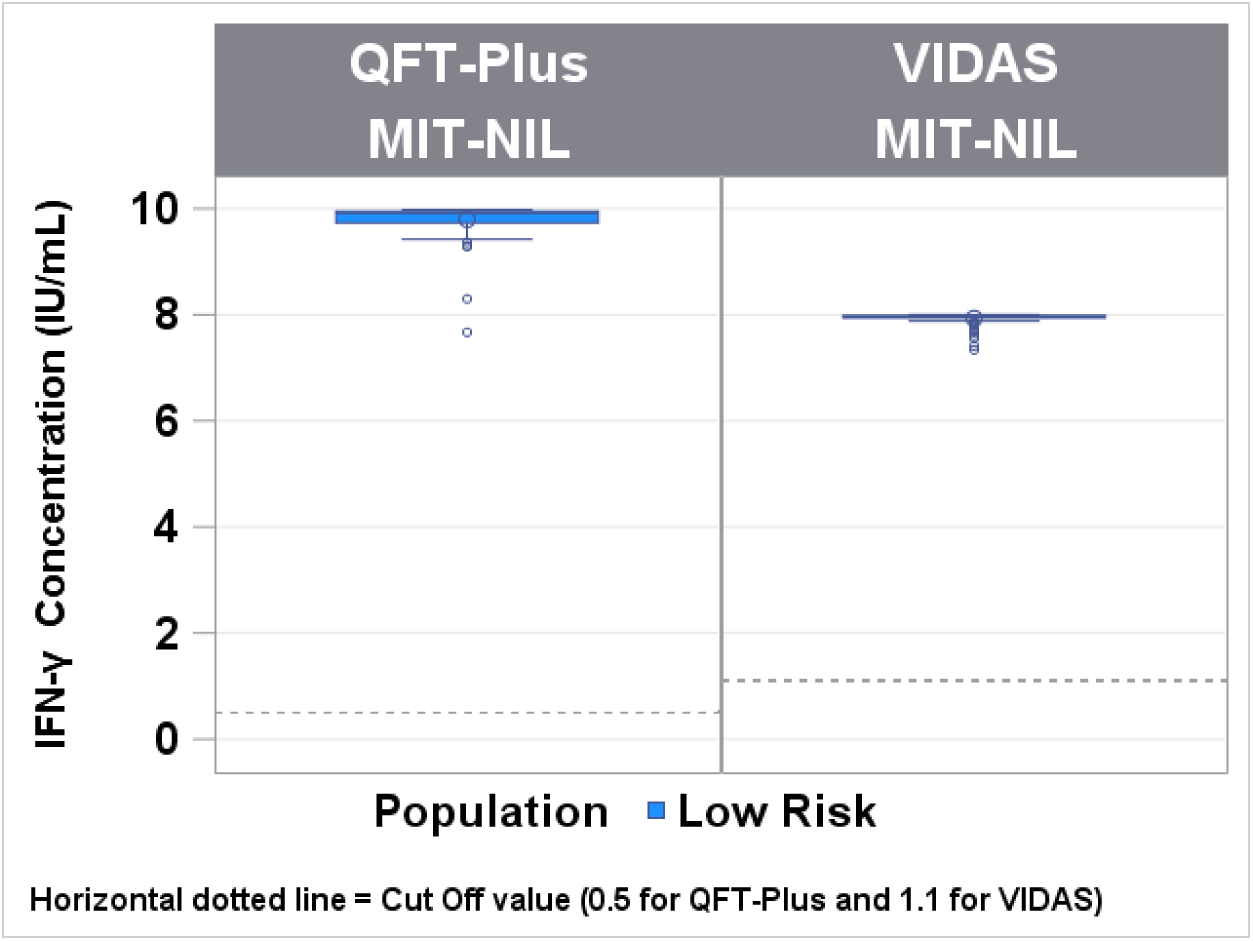
Comparison between the VIDAS® TB-IGRA and QFT-Plus for the low-risk population. **A.** Specificity estimates for the two assays. **B.** Sample numbers and negative predictive agreement (NPA). **C.** Patient sample interferon-γ (IFN-γ) concentrations for the VIDAS^®^ TB-IGRA and QFT-Plus. Nil, QFT-Plus negative control; NIL, VIDAS^®^ TB-IGRA negative control; Mitogen, QFT-Plus positive control; MIT-NIL, VIDAS® TB-IGRA positive control; TB1/TB2, QFT-Plus mitogenic response samples; AG-NIL, VIDAS^®^ TB-IGRA mitogenic response sample. The dotted lines represent the sample threshold values for the respective assay.

We also observed a strong NPA of 98.2% between the VIDAS® TB-IGRA and QFT-Plus, further indicating that the two tests exhibited similar diagnostic performance in this low-risk population (Figure 4B).

Furthermore, the VIDAS® TB-IGRA detected low basal IFN-γ values in the AG-NIL samples, suggesting that the system maintained specificity despite an improved detection capacity for TB infection (Figure 4C).

## DISCUSSION

Here, we provide the first evaluation of the diagnostic performance of the VIDAS® TB-IGRA in detecting both TB infection and disease in three patient populations compared with the widely used QFT-Plus. In the TB disease population, the VIDAS® TB-IGRA exhibited high sensitivity compared with international gold standard methods. Furthermore, the VIDAS® TB-IGRA outperformed the QFT-Plus by exhibiting higher sensitivity. The enhanced sensitivity of the VIDAS® TB-IGRA was most likely due to its capacity to provide a positive interpretation in samples that had an indeterminate or false negative result with the QFT-Plus. This difference in sensitivity could be further explained by the observation that the VIDAS® TB-IGRA detected higher sample IFN-γ values.

The QFT-Plus exhibited a lower sensitivity than previously indicated by the manufacturer and also yielded a large number of indeterminate results^12^. These indeterminate results were most likely not due to technical issues because all hospital personnel were thoroughly trained to ensure that all procedures were performed according to the manufacturer’s recommended guidelines and instructions.

Additionally, the proportion of indeterminate results was similar at two independent study sites, and calibration curves were carefully monitored to ensure measurement precision. Furthermore, correct sample processing was validated because the TB disease and high-risk samples were handled similarly and were simultaneously assayed in the same microplate, and no indeterminate results were obtained for the high-risk population with the QFT-Plus.

The low sensitivity of the QFT-Plus in detecting TB disease cases has been well documented^13-15^, particularly in countries with a high prevalence of *M. tuberculosis* infection and other comorbidities^16,17^, such as Burkina Faso, which was one of our study sites. Additionally, there may be a potential correlation between the low sensitivity of the QFT-Plus and a high frequency of comorbidities, particularly parasitic infections and fecal yeast^18-20^, in these patients. Because the sensitivity of the QFT-Plus has been previously determined in countries with a low prevalence of *M. tuberculosis* infection and other comorbidities, including the US, Japan, and Australia^12^, the low sensitivity and high number of indeterminate results obtained with the QFT-Plus in this study may be due to the particularly poor health status of the Burkina Faso study participants. However, this does not seem to have an effect on the VIDAS® TB-IGRA results. Further investigations are needed to compare the performance of the two tests in various geographical locations and under different contexts.

It is difficult to evaluate the capacities of the IGRAs to detect TB infection without a gold standard method for reference. However, in the high-risk population, the VIDAS® TB-IGRA demonstrated a high PPA with the QFT-Plus results, indicating its comparable performance in detecting TB infection in this patient group. The NPA was lower than expected in this population, but the NPA in correlation to risk stratification for *M. tuberculosis* exposure may indicate an increased capacity of the VIDAS® TB-IGRA in detecting marginal cases in this high-risk demographic.

In the low-risk population, the VIDAS® TB-IGRA exhibited a high NPA with the QFT-Plus results, thus demonstrating that the two tests had a similar performance in correctly identifying negative samples. Additionally, the VIDAS® TB-IGRA detected low basal IFN-γ values in the AG-NIL samples, further indicating that the specificity of the test was maintained despite its improved detection capacity.

Our study had some limitations. For example, the number of participants was limited, and all participants in the low-risk population were derived from a single location. Additional multicenter, prospective studies are warranted to validate our results.

In conclusion, the VIDAS® TB-IGRA exhibits higher sensitivity than a currently used IGRA while maintaining comparable specificity. In addition, its fully automated system allows for quick and accurate delivery of single-patient results. These characteristics point to the VIDAS® TB-IGRA as a novel, improved means for the use of IGRAs for TB diagnostics, which may contribute to the progress toward the successful eradication of TB worldwide.

## Data Availability

All data produced in the present study are available upon reasonable request to the authors

## ACKNOWLEDGMENTS

This work was supported by bioMérieux and an Afrique One-ASPIRE Fellowship funded by Wellcome Trust (WT087535MA) and awarded to Arthur D. Djibougou (107753/À/15/Z). We are grateful to all the study participants and the Etablissement Français du Sang, notably Yves Mérieux, for providing samples. We thank all the members of R&D Immunoassay bioMérieux who took part in this work. We are grateful to the bioMérieux Data Sciences team for the calculations and statistical evaluation of the data and to Victor Bondanese, Stéphanie Pascual, Laurence Bridon, and the Clinical Affairs team of bioMérieux for their help and technical support. We acknowledge Valérie Andrianasolonirina, Véronique Bourdais, Claire Pernin, and Lydia Suarez from Pf. Cambau’s team for their contribution to the work. The authors thank Kelsey Powell (BioScience Writers, USA) for providing medical writing support, which was funded by bioMérieux (Marcy L’Etoile, France) in accordance with Good Publication Practice (GPP3) guidelines (http://www.ismpp.org/gpp3).

## AUTHOR CONTRIBUTIONS

bioMerieux conceived the study; R&D from bioMerieux in collaboration with the teams of Pf. Gennaro, Pf. Diagbouga, and Pf. Cambau collected patient samples, optimized the study, and performed the serological assays. bioMerieux dataScience interpreted and analyzed the data of the serological assays. All authors contributed to the data interpretation and reviewed and approved the final manuscript.

