## supplemental data for "Preliminary diagnostic performance of the VIDAS® TB-IGRA for the detection of *Mycobacterium tuberculosis* infection and disease"

**SUPPLEMENTAL MATERIAL**

**SUPPLEMENTAL RESULTS**

**Analysis of CD4^+^ T-cell counts in patient samples from the TB disease population in Burkina Faso**

Low TB1/2 antigen and MIT-NIL IFN-γ responses resulted in indeterminate QFT-Plus results in the TB disease population; therefore, the number of CD4^+^ T cells was analyzed in the TB disease patient samples from the Burkina Faso location. Higher IFN-γ values in the AG-NIL and MIT-NIL samples were detected with the VIDAS**®** TB-IGRA compared with the QFT-Plus TB1/2 and MIT-NIL samples, respectively, regardless of CD4^+^ T-cell counts (Supplemental Figure 2). Therefore, no correlation between the number of CD4^+^ T cells present in the patient samples and the extent of the IFN-γ responses was observed, suggesting that the higher sensitivity of the VIDAS**®** TB-IGRA was most likely due to design optimization rather than the number of CD4^+^ T cells in the patient samples.

**Characteristics of Burkina Faso patients with TB disease**

Various characteristics were observed in the patients from the Burkina Faso location. Specifically, there was a high presence of parasitic infections and fecal yeast in these patients. Of the patients with TB disease, approximately 25% had parasitic infections and approximately 52% had yeast in their fecal samples. Likewise, approximately 21% of the high-risk patients had parasitic infections and approximately 59% exhibited yeast in their fecal samples. Additionally, nearly half (49%) of the Burkina Faso patients with TB disease had low BMIs (<18.5 kg/m^2^), possibly indicating malnutrition. Thus, the low sensitivity of the QFT-Plus in detecting TB infection and disease in these particular patient demographics may be at least partially attributed to the poor health status of these patients.

**Discrepant results in the high-risk population**

There were eight discrepant results between the VIDAS**®** TB-IGRA and QFT-Plus in the high-risk samples from the Newark, NJ location. These samples included 2 contact cases, 5 immigrants from high-prevalence countries, and 1 healthcare worker with an increased risk of exposure to *M. tuberculosis*. The VIDAS**®** TB-IGRA identified these eight samples as positive, whereas the QFT-Plus identified them as negative (Supplemental Table 3).

There were 11 discrepant results between the two assays in the high-risk samples from the Burkina Faso location. All of these samples were contact cases. The VIDAS**®** TB-IGRA identified eight of these samples as positive and three as negative, whereas the opposite results were obtained with the QFT-Plus (Supplemental Table 3).

Because all these individuals were at an increased risk of exposure and therefore had a higher chance of becoming infected with *M. tuberculosis*, these discrepant results partially demonstrated the improved capacity of the VIDAS**®** TB-IGRA to detect TB infection and disease in this high-risk group.

**Supplemental Figure 1.** Patient study inclusion criteria.

| TB disease | 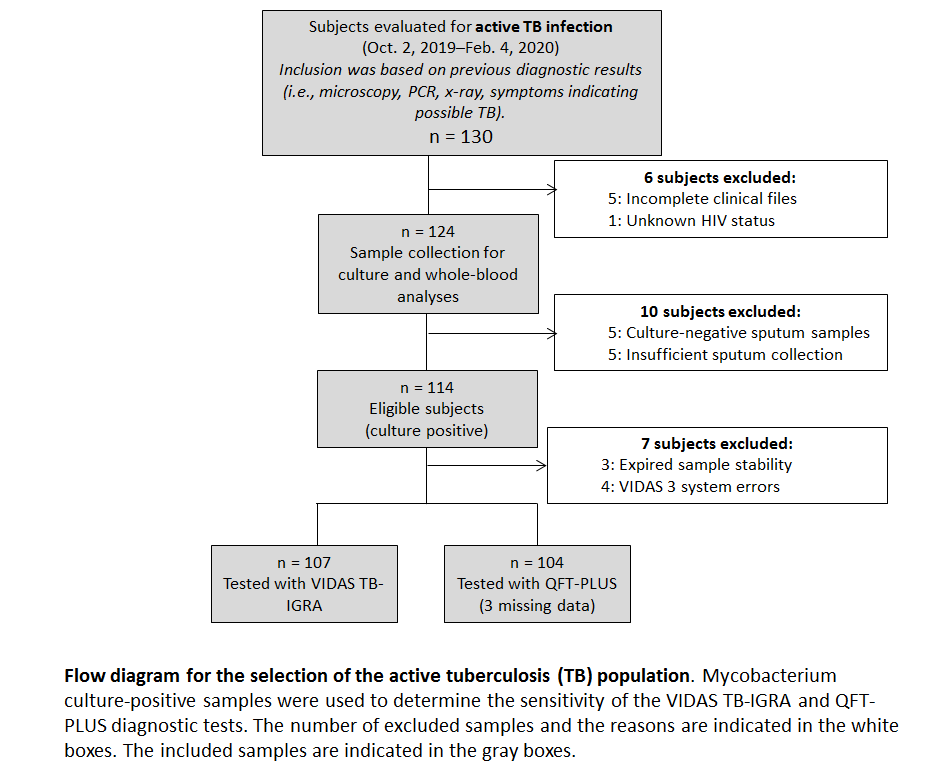 |
| --- | --- |
| High-risk | 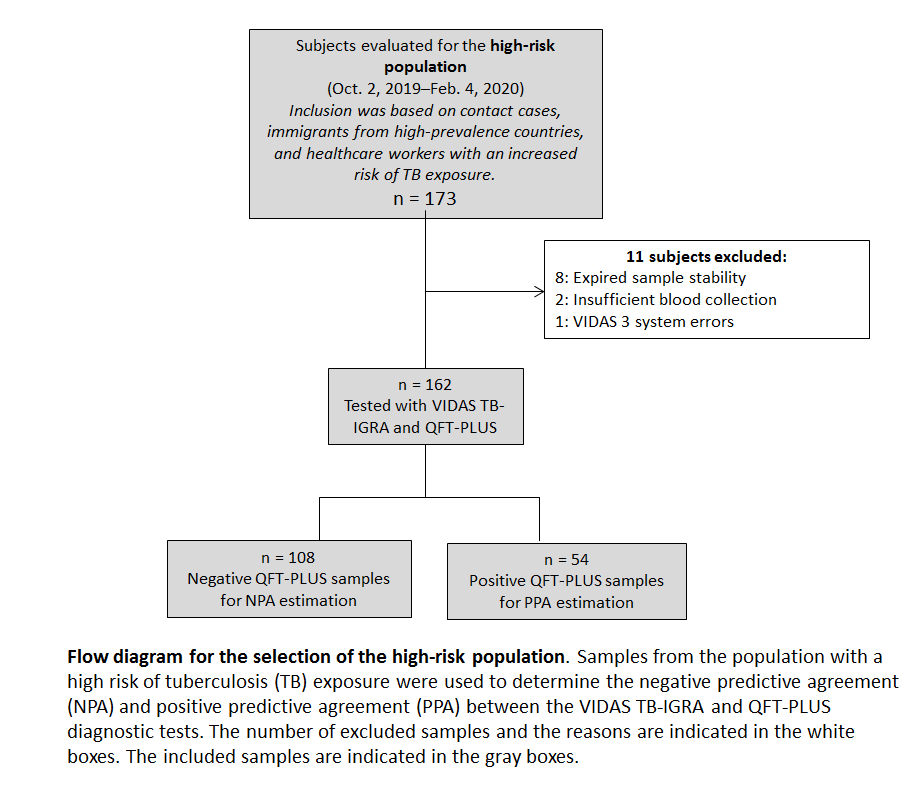 |
| Low-risk | 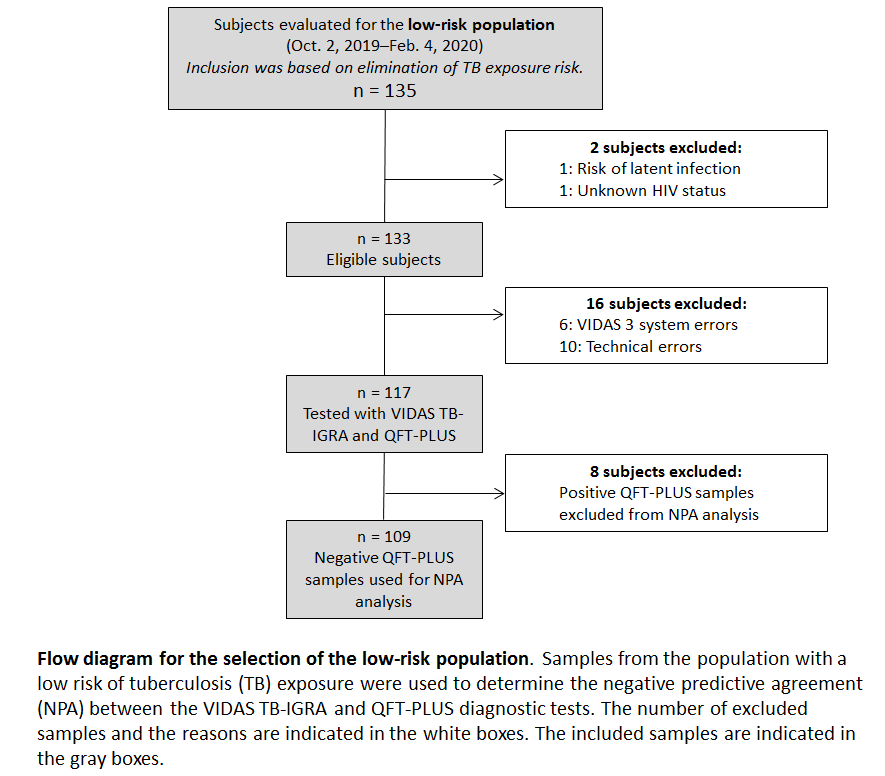 |

**Supplemental Figure 2.** Analysis of CD4^+^ T-cell densities in patient samples from the tuberculosis disease population in Burkina Faso. **A.** Interferon-γ (IFN-γ) concentrations in patient samples stratified by CD4^+^ T-cell numbers. **B.** Percentage of patients with tuberculosis disease according to CD4^+^ T-cell numbers.

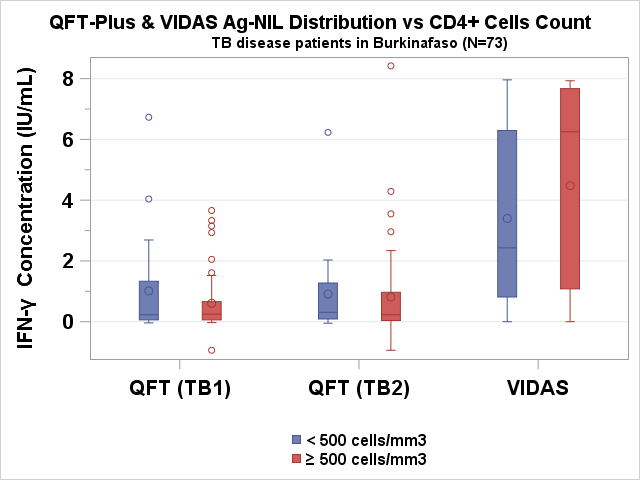

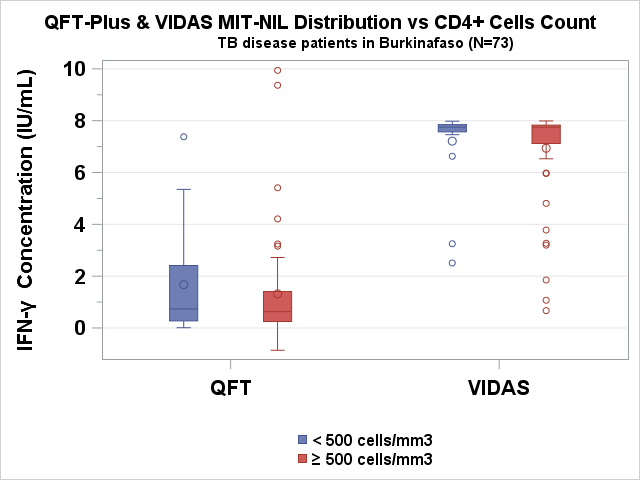

| Population recruited | Tuberculosis disease (Burkina Faso) |
| --- | --- |
| Study population, N (%) | 101 (100%) |
| Level of CD4^+^ T cells, N (%) |  |
| <500 cells/mm^3^ | 20 (19.8%) |
| ≥500 cells/mm^3^ | 53 (52.5%) |
| Unknown | 28 (27.7%) |

**SUPPLEMENTAL TABLES**

**Supplemental Table 1.** Demographics and additional information collected from patients included in the study.

| **Populations recruited** | **Low-risk** | **High-risk** | **Tuberculosis disease** |
| --- | --- | --- | --- |
| Study population, N (%) | 117 (100%) | 162 (100%) | 107 (100%) |
| Age in years, median (range) | 36 (18–67) | 37 (6–73) | 37 (13–82) |
| Sex, N (%) |  |  |  |
| Male | 70 (59.8%) | 73 (45.1%) | 86 (80.4%) |
| Female | 47 (40.2%) | 89 (54.9%) | 21 (19.6%) |
| Number provided by location, N (%) |  |  |  |
| Ouagadougou, Burkina Faso | - | 90 (55.5%) | 101 (94.4%) |
| Newark, NJ, USA | - | 72 (44.5%) | 4 (3.7%) |
| Paris, France | - | - | 2 (1.9%) |
| Lyon, France | 117 (100%) | - | - |
| Race, N (%) |  |  |  |
| Ouagadougou, Burkina Faso | - | 90 (56%) Africans and | 101 (94.4%) Africans |
| Newark, NJ, USA | - | 72 (44%) Americans, including 9 Asians, 21 Blacks/African Americans, and 42 Whites | 4 (3.7 %) Americans, including 3 Whites and 1 Black/African American |
| Paris, France | - | - | 2 (1.9%) Africans |
| Lyon, France | - | - | - |
| Ethnicity (US ONLY) | - | 72 (44%) Americans, including 42 Hispanics/Latinos and 30 Non-Hispanics/Latinos | 4 (4%) Americans, including 3 Hispanics/Latinos and 1 Non-Hispanic/Latino |
| Location of tuberculosis disease, N (%) |  |  |  |
| Pulmonary | - | - | 104 (97.2%) |
| Extra-pulmonary | - | - | 3* (2.8%) |
| ** 1 Bone, 1 cervical, 1 nodular* |  |  |  |
| HIV status, N (%) |  |  |  |
| Positive | - | 2 (1%) | - |
| Negative | 117 (100%) | 100 (62%) | 107 (100%) |
| Unknown | - | 60 (37%) | - |
| BCG vaccination, N (%) |  |  |  |
| Yes | - | 114 (70%) | 61 (57%) |
| No | - | 35 (22%) | 8 (7.5%) |
| Unknown | 117 (100%) | 13 (8%) | 38 (35.5%) |
| Diabetes, N (%) |  |  |  |
| Yes | - | 8 (5%) | 1 (1%) |
| No | 117 (100%) | 154 (95%) | 106 (99%) |
| Autoimmune diseases, N (%) |  |  |  |
| Yes | - | 3 (2%) | 1 (1%) |
| No | 117 (100%) | 159 (98%) | 106 (99%) |
| Infectious diseases, N (%) |  |  |  |
| Yes | - | 2 ( 1%) | 3 (3%) |
| No | 117 (100%) | 160 (99%) | 104 (97%) |
| Other pathologies, N (%) |  |  |  |
| Yes | - | 24 (15%) | 13 (12%) |
| No | 117 (100%) | 138 (85%) | 94 (88%) |
| Antibiotic treatments, N (%) |  |  |  |
| Yes | - | 3 (2%) | 106 (99%) |
| No | 117 (100%) | 159 (98%) | 1 (1%) |
| Immunosuppressive treatments, N (%) |  |  |  |
| Yes | - | 1 (1%) | - |
| No | 117 (100%) | 161 (99%) | 107 (100%) |
| Pain treatment (analgesics), N (%) |  |  |  |
| Yes |  | 5 (3%) | 11 (10.3%) |
| No | 117 (100%) | 157 (97%) | 96 (89.7%) |
| Other treatments, N (%) |  |  |  |
| Yes | - | 21 (13%) | 14 (13%) |
| No | 117 (100%) | 141 (87%) | 93 (87%) |
| TST, N (%) |  |  |  |
| Yes | - | 35 (22%) | 1 (1%) |
| No | 18 (15%) | 123 (76%) | 103 (96%) |
| Unknown | 99 (85%) | 4 (2%) | 3 (3%) |

**Supplemental Table 2.** Discrepant results in the high-risk population.

| **Sample ID** | **Clinical context** | **TB exposure risk** | **VIDAS TB-IGRA** | | |  | **QFT-Plus** | | | | |
| --- | --- | --- | --- | --- | --- | --- | --- | --- | --- | --- | --- |
|  |  |  | **NIL** | **Ag-NIL** | **MIT-NIL** | **Interp.** | **NIL** | **TB1-NIL** | **TB2-NIL** | **MIT-NIL** | **Interp.** |
| **12** | **Healthcare worker** | **Medium** | 0.14 | 0.40 | 7.86 | P | 0.25 | 0.32 | 0.33 | 9.75 | N |
| **7** | **Immigrant from country with high incidence of TB** | **Medium high** | 0.21 | 0.75 | 7.79 | P | 0.5 | -0.11 | -0.03 | 9.5 | N |
| **3** | **Immigrant from country with high incidence of TB** | **Medium high** | 0.02 | 1.33 | 7.98 | P | 0.25 | 0.03 | 0.06 | 9.75 | N |
| **18** | **Immigrant from country with high incidence of TB** | **Medium high** | 0.19 | 3.55 | 7.81 | P | 0.21 | 0.22 | 0.25 | 9.79 | N |
| **9** | **Immigrant from country with high incidence of TB** | **Medium high** | 0.02 | 0.57 | 7.98 | P | 0.12 | 0.18 | 0.21 | 9.88 | N |
| **19** | **Contact with sick person** | **High** | 0.11 | 0.59 | 7.89 | P | 0.22 | -0.07 | -0.08 | 9.78 | N |
| **2** | **Contact with sick person** | **High** | 0.55 | 3.02 | 7.45 | P | 0.03 | 0.08 | 0.15 | 9.97 | N |
| **16** | **Contact with sick person within household** | **Very high** | 0.16 | 1.18 | 7.84 | P | 0.07 | 0.15 | 0.34 | 9.93 | N |
| **6** | **Contact with sick person within household** | **Very high** | 0.15 | 1.43 | 7.85 | P | 0.06 | -0.02 | 0.04 | 9.94 | N |
| **10** | **Contact with sick person within household** | **Very high** | 0.21 | 0.41 | 7.79 | P | 0.06 | 0.10 | 0.04 | 9.94 | N |
| **13** | **Contact with sick person within household** | **Very high** | 0.22 | 0.45 | 7.78 | P | 0.03 | 0.02 | 0.00 | 9.97 | N |
| **4** | **Contact with sick person within household** | **Very high** | 0.30 | 0.58 | 7.70 | P | 0.07 | 0.12 | 0.06 | 6.9 | N |
| **14** | **Contact with sick person within household** | **Very high** | 0.53 | 0.43 | 7.47 | P | 0.12 | 0.04 | 0.01 | 3.16 | N |
| **15** | **Contact with sick person within household** | **Very high** | 0.06 | 1.20 | 7.94 | P | 0,02 | 0.22 | 0.09 | 3,52 | N |
| **5** | **Contact with sick person within household** | **Very high** | 0.07 | 0.74 | 7.93 | P | 0,03 | 0.07 | 0.08 | 7,64 | N |
| **17** | **Contact with sick person within household** | **Very high** | 0.08 | 1.59 | 7.92 | P | 0,14 | 0.23 | 0.26 | 9,86 | N |
| **8** | **Contact with sick person within household** | **Very high** | 0.29 | 0.26 | 7.71 | N | 0.06 | 1.09 | 0.97 | 9.5 | P |
| **11** | **Contact with sick person within household** | **Very high** | 0.05 | 0.04 | 7.95 | N | 0.02 | 0.88 | 1.07 | 2.6 | P |
| **1** | **Contact with sick person within household** | **Very high** | 0.08 | 0.32 | 7.92 | N | 0.05 | 0.43 | 0.37 | 8.86 | P |
